# Characterizing the incidence of adverse events of special interest for COVID-19 vaccines across eight countries: a multinational network cohort study

**DOI:** 10.1101/2021.03.25.21254315

**Authors:** Xintong Li, Anna Ostropolets, Rupa Makadia, Azza Shaoibi, Gowtham Rao, Anthony G. Sena, Eugenia Martinez-Hernandez, Antonella Delmestri, Katia Verhamme, Peter R Rijnbeek, Talita Duarte-Salles, Marc Suchard, Patrick Ryan, George Hripcsak, Daniel Prieto-Alhambra

## Abstract

**Background:** As large-scale immunization programs against COVID-19 proceed around the world, safety signals will emerge that need rapid evaluation. We report population-based, age- and sex-specific background incidence rates of potential adverse events of special interest (AESI) in eight countries using thirteen databases.

**Methods:** This multi-national network cohort study included eight electronic medical record and five administrative claims databases from Australia, France, Germany, Japan, Netherlands, Spain, the United Kingdom, and the United States, mapped to a common data model. People observed for at least 365 days before 1 January 2017, 2018, or 2019 were included. We based study outcomes on lists published by regulators: acute myocardial infarction, anaphylaxis, appendicitis, Bell’s palsy, deep vein thrombosis, disseminated intravascular coagulation, encephalomyelitis, Guillain-Barre syndrome, hemorrhagic and non-hemorrhagic stroke, immune thrombocytopenia, myocarditis/pericarditis, narcolepsy, pulmonary embolism, and transverse myelitis. We calculated incidence rates stratified by age, sex, and database. We pooled rates across databases using random effects meta-analyses. We classified meta-analytic estimates into Council of International Organizations of Medical Sciences categories: very common, common, uncommon, rare, or very rare.

**Findings:** We analysed 126,661,070 people. Rates varied greatly between databases and by age and sex. Some AESI (e.g., myocardial infarction, Guillain-Barre syndrome) increased with age, while others (e.g., anaphylaxis, appendicitis) were more common in young people. As a result, AESI were classified differently according to age. For example, myocardial infarction was very rare in children, rare in women aged 35-54 years, uncommon in men and women aged 55-84 years, and common in those aged ≥85 years.

**Interpretation:** We report robust baseline rates of prioritised AESI across 13 databases. Age, sex, and variation between databases should be considered if background AESI rates are compared to event rates observed with COVID-19 vaccines.

## Introduction

On 11 March 2020, the World Health Organization (WHO) declared the outbreak of COVID-19, caused by the SARS-CoV-2 virus, a global pandemic. As of March 2021, over 100 million confirmed cases and 2·7 million deaths have been reported worldwide.^1^ Vaccines for COVID-19 have been developed at unprecedented speed, with phase 3 clinical efficacy trials reporting resulted for some vaccines less than a year after the WHO declared the pandemic. Several vaccines have been authorised by regulators since December 2020, such as the European Medicines Agency (EMA), the Food and Drug Administration (FDA) in the US, and the UK Medicines and Healthcare Products Regulatory Agency (MHRA). Large-scale immunization programs are ongoing worldwide.

While this scientific achievement should be celebrated, we must recognize that emergency use is accompanied by residual uncertainty about the safety and effectiveness of vaccines in all populations of interest. As with all medical products reaching the milestone of regulatory authorization, vaccine safety must continue to be monitored to complement what was initially learned during clinical development. Spontaneous adverse event reporting has served as a foundational component of post-approval pharmacovigilance activities to ensure the safe and appropriate use of medical products. Observational healthcare data captured during the routine course of clinical care, such as electronic health records and administrative claims, can augment pharmacovigilance by providing real-world context about potential adverse events and their rates in populations of interest. Background rates of adverse events have historically played an important role in monitoring the safety of vaccines by serving as a baseline comparator for observed rates among those vaccinated.^2,3^ Each new vaccine has potential adverse events of special interest (AESI) that warrant focused evaluation, based on historical precedent set by prior vaccines and knowledge acquired during its development.

As COVID-19 vaccines have received authorization for emergency use, regulatory agencies around the world have been preparing safety surveillance strategies. The US FDA Center for Biologics Evaluation and Research published a protocol on “ Background Rates of Adverse Events of Special Interest for COVID-19 Vaccine Safety Monitoring.” ^4^ The European Medicines Agency-funded vACCine covid-19 monitoring readinESS (ACCESS) project also included estimation of background AESI rates in their protocol.^5^ Recognizing the global burden that COVID-19 represents and following the WHO Council for International Organizations of Medical Sciences (CIOMS) guidance that the most valid data for comparison in a particular area are the background rates from the local population,^6^ the Observational Health Data Sciences and Informatics (OHDSI) community collaborated to design and execute an international open science study to characterize background rates of COVID-19 AESI. In this paper, we provide descriptive epidemiology context for COVID-19 AESIs using observational data from Australia, France, Germany, Japan, Netherlands, Spain, the UK, and the US.

## Methods

### Study design

A multinational, multi-database population-based network cohort study.

### Data sources

We included thirteen databases from eight countries, of which eight were electronic health record data sources and five were administrative claims data sources.

The electronic health record databases were: 1. IQVIA Australia Electronic Medical Records (IQVIA_AUSTRALIA); 2. Integrated Primary Care Information (IPCI_NETHERLANDS), a primary care records database from the Netherlands; 3. IQVIA Longitudinal Patient Data France (IQVIA_FRANCE); 4. IQVIA Disease Analyser Germany (IQVIA_GERMANY); 5. Information System for Research in Primary Care (SIDIAP_H_SPAIN), a primary care records database that covers over 80% of the population of Catalonia, Spain; 6. Clinical Practice Research Datalink, which consists of data collected from UK primary care for all ages (CPRD_GOLD_UK); 7. Columbia University Irving Medical Center (CUMC_US), which covers the New York-Presbyterian Hospital/Columbia University Irving Medical Center in the US; and 8. Optum^®^ de-identified Electronic Health Record Dataset (OPTUM_EHR_US), which covers more than 103 million patients and over 7,000 hospitals and clinics across the US.

The claims-based databases were the Japan Medical Data Center (JMDC_JAPAN) and four US administrative claims databases: IBM MarketScan Commercial Claims and Encounters Database (CCAE_US), IBM MarketScan Medicare Supplemental and Coordination of Benefits Database (MDCR_US), IBM MarketScan Multi-State Medicaid Database (MDCD_US), Optum^®^ De-Identified Clinformatics^®^ Data Mart Database – Socio-Economic Status (OPTUM_SES_US).

A detailed description of the databases can be found in Appendix Table 1.

**Table 1:** Demographics of the included populations, stratified by database.

|  | CCAE_US | MDCD_US | MDCR_US | OPTUM_EHR_US | OPTUM_SES_US | CUMC_US | CPRD_GOLD_UK | IPCI_NETHERLANDS | SIDIAP_H_SPAIN | IQVIA_FRANCE | IQVIA_GERMANY | IQVIA_AUSTRALIA | JMDC_JAPAN |
| --- | --- | --- | --- | --- | --- | --- | --- | --- | --- | --- | --- | --- | --- |
| Total patients | 25,315,777 | 12,966,011 | 1,533,709 | 40,955,085 | 18,643,608 | 1,164,196 | 4,532,766 | 1,536,283 | 2,217,536 | 1,746,371 | 9,295,525 | 252,212 | 6,501,991 |
| Person-years | 42,889,550 | 23,203,712 | 2,484,782 | 72,328,897 | 32,474,685 | 2,174,312 | 9,638,136 | 3,326,570 | 5,497,613 | 3,008,350 | 16,784,613 | 383,668 | 12,848,482 |
| Age |  |  |  |  |  |  |  |  |  |  |  |  |  |
| 1 - 5 | 4.96% | 13.54% | 0.0% | 4.52% | 3.36% | 3.49% | 5.42% | 5.13% | 4.5% | 5.69% | 3.32% | 5.32% | 6.37% |
| 6 - 17 | 16.28% | 32.3% | 0.0% | 11.65% | 10.36% | 9.06% | 14.01% | 13.74% | 11.73% | 15.38% | 8.86% | 12.6% | 16.06% |
| 18 - 34 | 25.26% | 22.26% | 0.0% | 19.98% | 17.87% | 17.1% | 20.87% | 19.85% | 16.91% | 18.83% | 15.19% | 20.22% | 23.59% |
| 35 - 54 | 31.98% | 15.48% | 0.0% | 26.22% | 23.54% | 25.84% | 26.86% | 25.7% | 29.92% | 25.58% | 25.16% | 27.7% | 35.84% |
| 55 - 64 | 18.63% | 7.75% | 0.0% | 16.25% | 12.79% | 15.77% | 13.11% | 14.32% | 13.01% | 13.11% | 17.0% | 14.4% | 13.54% |
| 65 - 74 | 2.88% | 4.88% | 47.8% | 11.79% | 16.66% | 14.77% | 10.36% | 11.75% | 11.13% | 11.33% | 13.76% | 10.81% | 4.3% |
| 75 - 84 | 0.0% | 2.63% | 35.01% | 6.48% | 10.65% | 9.52% | 6.4% | 6.79% | 8.16% | 6.7% | 12.82% | 6.07% | 0.32% |
| 85+ | 0.0% | 1.16% | 17.19% | 3.11% | 4.77% | 4.44% | 2.96% | 2.71% | 4.64% | 3.38% | 3.9% | 2.86% | 0.0% |
| Sex |  |  |  |  |  |  |  |  |  |  |  |  |  |
| Female | 51.5% | 56.47% | 55.38% | 56.7% | 51.47% | 59.54% | 50.47% | 51.01% | 50.52% | 53.03% | 57.45% | 54.4% | 45.01% |
| Male | 48.5% | 43.53% | 44.62% | 43.3% | 48.53% | 40.46% | 49.53% | 48.99% | 49.48% | 46.97% | 42.55% | 45.6% | 54.99% |

\*CCAE\_US: IBM MarketScan Commercial Claims and Encounters Database, CPRD\_GOLD\_UK: Clinical Practice Research Datalink, CUMC\_US: Columbia University Irving Medical Center, IPCI\_NETHERLANDS: Integrated Primary Care Information, IQVIA\_AUSTRALIA: IQVIA Australia Electronic Medical Records, IQVIA\_FRANCE: IQVIA Longitudinal Patient Data France, IQVIA\_GERMANY: IQVIA Disease Analyser Germany, JMDC\_JAPAN: Japan Medical Data Center, MDCD\_US: IBM MarketScan Multi-State Medicaid Database, MDCR\_US: IBM MarketScan Medicare Supplemental and Coordination of Benefits Database, OPTUM\_EHR\_US: Optum® de-identified Electronic Health Record Dataset, OPTUM\_SES\_US: Optum® De-Identified Clinformatics® Data Mart Database – Socio-Economic Status, SIDIAP\_H\_SPAIN: Information System for Research in Primary Care – Hospitalization Linked Data

All datasets were previously mapped to the Observational Medical Outcomes Partnership common data model, which is maintained by the Observational Health Data Sciences and Informatics (OHDSI) network. The analysis code could therefore be distributed across all contributing centers without sharing patient-level data.^7,8^

### Population/study participants

In our primary analysis, we defined the target at-risk population as people who were observed on 1 January 2017, 1 January 2018, or 1 January 2019 and were observed for at least 365 days before this observation date. We defined 1 January each year as the index date.

### Events of interest

The events of interest in this study were the AESIs that might need evaluation following COVID-19 vaccination. This outcome list was developed primarily based on the “ Background Rates of Adverse Events of Special Interest for COVID-19 Vaccine Safety Monitoring” protocol published by the FDA Center for Biologics Evaluation and Research, the prioritiszd COVID-19 vaccine AESI list by the Brighton Collaboration, and other previous studies.^4,9^ Fifteen events were included: non-hemorrhagic stroke, hemorrhagic stroke, acute myocardial infarction, deep vein thrombosis, pulmonary embolism, anaphylaxis, Bell’s palsy, myocarditis/pericarditis, narcolepsy, appendicitis, immune thrombocytopenia, disseminated intravascular coagulation, encephalomyelitis (including acute disseminated encephalomyelitis), Guillain-Barre syndrome, and transverse myelitis.^4^

Events were identified by condition occurrence records (e.g., diagnosis codes from claims or diagnosis codes and problem lists from electronic health records). Encephalomyelitis, non-hemorrhagic stroke, hemorrhagic stroke, and acute myocardial infarction definitions also required the record to occur within an inpatient setting (any position), while the Guillain-Barre syndrome definition required the condition to be recorded in an inpatient setting in the primary position.

Qualifying events could not be previously observed in a ‘clean window’ period before the index date. In keeping with the FDA protocol, we applied a clean window of 365 days for all events except anaphylaxis (30 days) and facial nerve palsy and encephalomyelitis (183 days).^4^ The full specifications of all phenotype definitions, including source codes and standard concepts, are available in Appendix Tables 2 and 3.

As the CPRD-GOLD (UK), IQVIA (France, Germany, and Australia), and IPCI (the Netherlands) databases only included primary care data, we did not use them for events whose definition required an inpatient diagnosis.

### Analysis

We defined the time-at-risk as a 365-day period following the index date. People contributed time-at-risk from 1 January to 31 December for each qualifying year in 2017 to 2019, but time was censored during the clean window following an event and at the end of a person’s observation period. One person could contribute more than one event, with outcome-specific pre-specified clean periods of 30 to 365 days used to avoid duplicate counts.

Incidence rates were estimated as the total number of events divided by the person-time at risk per 100,000 person-years. We calculated the age-by-sex specific incidence rates in each database and report all rates where the event counts exceeded a minimum cell count of 5. Age was calculated as year of index date minus year of birth and was partitioned into eight mutually exclusive age groups: 1-5 years old, 6-17, 18-35, 36-55, 56-64, 65-74, 75-84, and 85 years and older. We then pooled age-sex specific rates of each AESI across all databases using random-effects models with the DerSimonian-Laird method to estimate between-database variance.^10^ We estimated 95% predicted intervals (95% confidence intervals) using the R package ‘meta’.^11^

Meta-analytic age and sex-specific rates were classified using the World Health Organization Council for International Organizations of Medical Sciences (CIOMS) thresholds: very common (≥1/10), common (<1/10 to ≥1/100), uncommon (<1/100 to ≥1/1,000), rare (<1/1,000 to ≥1/10,000), and very rare (<1/10,000).^12^

All statistical analyses were performed in R software.^13^ The study protocol and analysis code are available at: https://github.com/ohdsi-studies/Covid19VaccineAesiIncidenceCharacterization

### Ethical approval

The protocol for this research was approved by the Independent Scientific Advisory Committee (ISAC) for MHRA Database Research (protocol number 20_000211), the IDIAPJGol Clinical Research Ethics Committee (project code: 21/007-PCV), the IPCI governance board (application number 3/2021), and the Columbia University Institutional Review Board (AAAO7805).

## Results

From 13 databases, 126,661,070 people contributed 227,043,370 person-years of follow-up. Each database captured important different population demographics, as shown in Table 1, and collectively represented all age and sex subgroups from eight countries.

Table 2 summarises the incidence rates of the 15 AESIs, stratified by age and sex, based on prediction intervals from a meta-analysis of the database estimates. Each age/sex subgroup was also classified using the CIOMS adverse event frequency system (very common, common, uncommon, rare, or very rare). The incidence of several outcomes varied substantially by age. For example, acute myocardial infarction was very rare (<1/10,000) in women under 35 years, rare (1/1,000 to 1/10,000) in women aged 35 to 54 years, uncommon (1/100 to 1/1,000) in both men and women aged 55 to 84 years, and common (1/10 to 1/100) in men and women aged 85 years and older. For both men and women, deep vein thrombosis was rare in those under 18 years, uncommon in those aged 35 to 84 years, and common in those 85 years and older. Women aged 18 to 34 years had a higher rate of deep vein thrombosis than men of the same age.

**Table 2.**
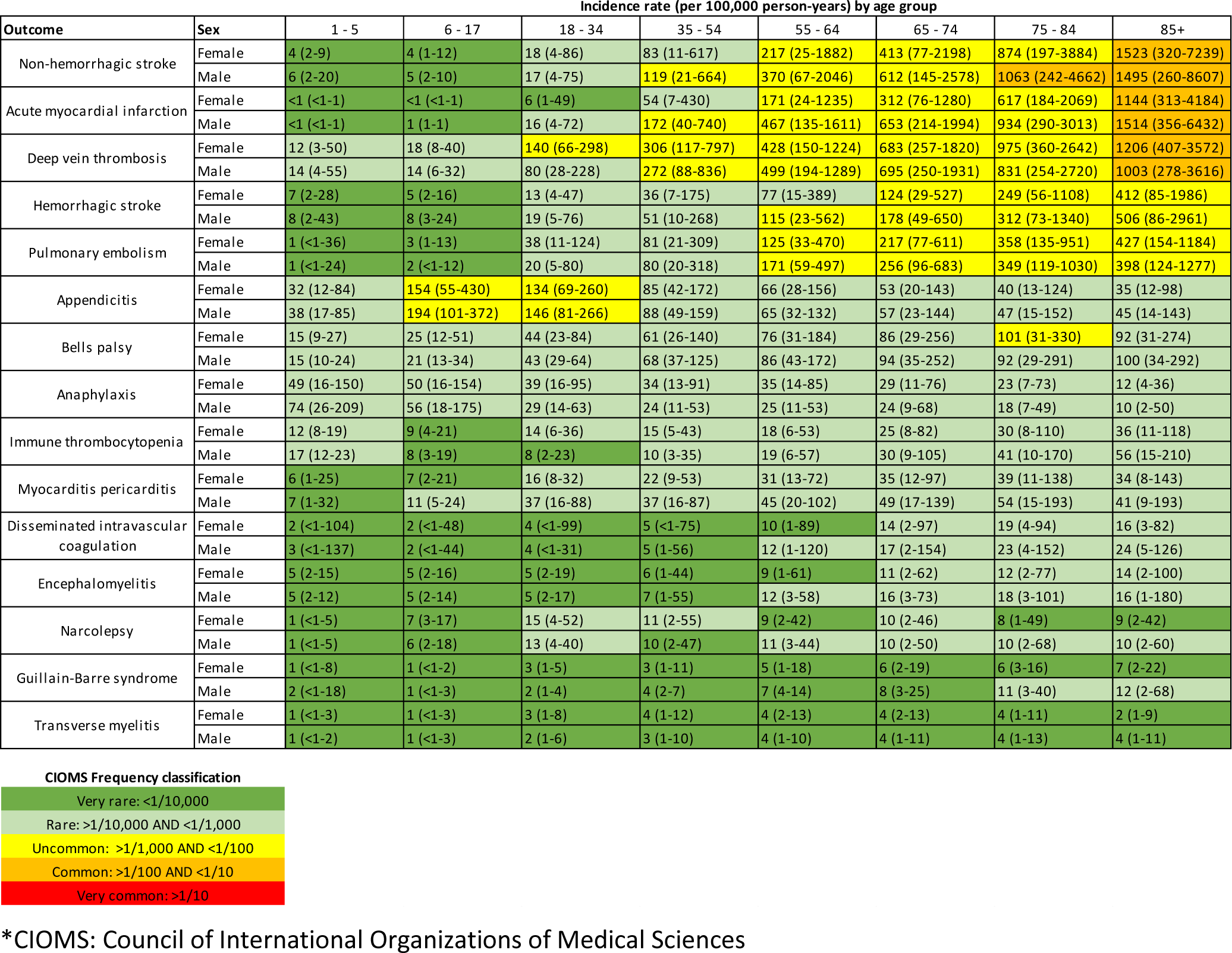
Pooled estimated age-sex stratified incidence rates per 100,00 person-years (with 95% confidence intervals), calculated from meta-analyses.

| Incidence rate (per 100,000 person-years) by age group |  |  |  |  |  |  |  |  |  |
| --- | --- | --- | --- | --- | --- | --- | --- | --- | --- |
| Outcome | Sex | 1 - 5 | 6 - 17 | 18 - 34 | 35 - 54 | 55 - 64 | 65 - 74 | 75 - 84 | 85+ |
| Non-hemorrhagic stroke | Female | 4 (2-9) | 4 (1-12) | 18 (4-86) | 83 (11-617) | 217 (25-1882) | 413 (77-2198) | 874 (197-3884) | 1523 (320-7239) |
|  | Male | 6 (2-20) | 5 (2-10) | 17 (4-75) | 119 (21-664) | 370 (67-2046) | 612 (145-2578) | 1063 (242-4662) | 1495 (260-8607) |
| Acute myocardial infarction | Female | <1 (<1-1) | <1 (<1-1) | 6 (1-49) | 54 (7-430) | 171 (24-1235) | 312 (76-1280) | 617 (184-2069) | 1144 (313-4184) |
|  | Male | <1 (<1-1) | 1 (1-1) | 16 (4-72) | 172 (40-740) | 467 (135-1611) | 653 (214-1994) | 934 (290-3013) | 1514 (356-6432) |
| Deep vein thrombosis | Female | 12 (3-50) | 18 (8-40) | 140 (66-298) | 306 (117-797) | 428 (150-1224) | 683 (257-1820) | 975 (360-2642) | 1206 (407-3572) |
|  | Male | 14 (4-55) | 14 (6-32) | 80 (28-228) | 272 (88-836) | 499 (194-1289) | 695 (250-1931) | 831 (254-2720) | 1003 (278-3616) |
| Hemorrhagic stroke | Female | 7 (2-28) | 5 (2-16) | 13 (4-47) | 36 (7-175) | 77 (15-389) | 124 (29-527) | 249 (56-1108) | 412 (85-1986) |
|  | Male | 8 (2-43) | 8 (3-24) | 19 (5-76) | 51 (10-268) | 115 (23-562) | 178 (49-650) | 312 (73-1340) | 506 (86-2961) |
| Pulmonary embolism | Female | 1 (<1-36) | 3 (1-13) | 38 (11-124) | 81 (21-309) | 125 (33-470) | 217 (77-611) | 358 (135-951) | 427 (154-1184) |
|  | Male | 1 (<1-24) | 2 (<1-12) | 20 (5-80) | 80 (20-318) | 171 (59-497) | 256 (96-683) | 349 (119-1030) | 398 (124-1277) |
| Appendicitis | Female | 32 (12-84) | 154 (55-430) | 134 (69-260) | 85 (42-172) | 66 (28-156) | 53 (20-143) | 40 (13-124) | 35 (12-98) |
|  | Male | 38 (17-85) | 194 (101-372) | 146 (81-266) | 88 (49-159) | 65 (32-132) | 57 (23-144) | 47 (15-152) | 45 (14-143) |
| Bells palsy | Female | 15 (9-27) | 25 (12-51) | 44 (23-84) | 61 (26-140) | 76 (31-184) | 86 (29-256) | 101 (31-330) | 92 (31-274) |
|  | Male | 15 (10-24) | 21 (13-34) | 43 (29-64) | 68 (37-125) | 86 (43-172) | 94 (35-252) | 92 (29-291) | 100 (34-292) |
| Anaphylaxis | Female | 49 (16-150) | 50 (16-154) | 39 (16-95) | 34 (13-91) | 35 (14-85) | 29 (11-76) | 23 (7-73) | 12 (4-36) |
|  | Male | 74 (26-209) | 56 (18-175) | 29 (14-63) | 24 (11-53) | 25 (11-53) | 24 (9-68) | 18 (7-49) | 10 (2-50) |
| Immune thrombocytopenia | Female | 12 (8-19) | 9 (4-21) | 14 (6-36) | 15 (5-43) | 18 (6-53) | 25 (8-82) | 30 (8-110) | 36 (11-118) |
|  | Male | 17 (12-23) | 8 (3-19) | 8 (2-23) | 10 (3-35) | 19 (6-57) | 30 (9-105) | 41 (10-170) | 56 (15-210) |
| Myocarditis pericarditis | Female | 6 (1-25) | 7 (2-21) | 16 (8-32) | 22 (9-53) | 31 (13-72) | 35 (12-97) | 39 (11-138) | 34 (8-143) |
|  | Male | 7 (1-32) | 11 (5-24) | 37 (16-88) | 37 (16-87) | 45 (20-102) | 49 (17-139) | 54 (15-193) | 41 (9-193) |
| Disseminated intravascular coagulation | Female | 2 (<1-104) | 2 (<1-48) | 4 (<1-99) | 5 (<1-75) | 10 (1-89) | 14 (2-97) | 19 (4-94) | 16 (3-82) |
|  | Male | 3 (<1-137) | 2 (<1-44) | 4 (<1-31) | 5 (1-56) | 12 (1-120) | 17 (2-154) | 23 (4-152) | 24 (5-126) |
| Encephalomyelitis | Female | 5 (2-15) | 5 (2-16) | 5 (2-19) | 6 (1-44) | 9 (1-61) | 11 (2-62) | 12 (2-77) | 14 (2-100) |
|  | Male | 5 (2-12) | 5 (2-14) | 5 (2-17) | 7 (1-55) | 12 (3-58) | 16 (3-73) | 18 (3-101) | 16 (1-180) |
| Narcolepsy | Female | 1 (<1-5) | 7 (3-17) | 15 (4-52) | 11 (2-55) | 9 (2-42) | 10 (2-46) | 8 (1-49) | 9 (2-42) |
|  | Male | 1 (<1-5) | 6 (2-18) | 13 (4-40) | 10 (2-47) | 11 (3-44) | 10 (2-50) | 10 (2-68) | 10 (2-60) |
| Guillain-Barre syndrome | Female | 1 (<1-8) | 1 (<1-2) | 3 (1-5) | 3 (1-11) | 5 (1-18) | 6 (2-19) | 6 (3-16) | 7 (2-22) |
|  | Male | 2 (<1-18) | 1 (<1-3) | 2 (1-4) | 4 (2-7) | 7 (4-14) | 8 (3-25) | 11 (3-40) | 12 (2-68) |
| Transverse myelitis | Female | 1 (<1-3) | 1 (<1-3) | 3 (1-8) | 4 (1-12) | 4 (2-13) | 4 (2-13) | 4 (1-11) | 2 (1-9) |
|  | Male | 1 (<1-2) | 1 (<1-3) | 2 (1-6) | 3 (1-10) | 4 (1-10) | 4 (1-11) | 4 (1-13) | 4 (1-11) |

**CIOMS Frequency classification**
|  |
| --- |
| Very rare: <1/10,000 |
| Rare: >1/10,000 AND <1/1,000 |
| Uncommon: >1/1,000 AND <1/100 |
| Common: >1/100 AND <1/10 |
| Very common: >1/10 |

\*CIOMS: Council of International Organizations of Medical Sciences

The AESIs studied spanned the continuum of possible frequencies. Non-hemorrhagic stroke, acute myocardial infarction, and deep vein thrombosis were all common in those aged 85 years and older and uncommon in those aged 55 to 74 years. Anaphylaxis, Bell’s palsy, appendicitis, and immune thrombocytopenia were largely rare in all age groups, although appendicitis was uncommon in those aged 6 to 34 years. Guillain-Barre syndrome and transverse myelitis were very rare in nearly all subgroups.

The prediction intervals for each age-sex subgroup were notably wide, reflecting the substantial population-level heterogeneity observed across sources. Figure 1 shows the database-specific incidence rates that were used to calculate the meta-analytic estimates. The rates recorded for deep vein thrombosis highlight this variation. For women aged 35 to 54 years, the incidence rates ranged from 159/100,000 person-years in Spain (SIDIAP) to 866/100,000 person-years in the US (MDCD). We obtained 13 database estimates for the incidence in women aged 65 to 74 years, ranging from 387/100,000 to 1,443/100,000 person-years. Eight databases gave rates under 650/100,000 person-years (CPRD-GOLD in the UK; CUIMC in the US; IPCI in the Netherlands; IQVIA in Australia, France, and Germany; JMDC in Japan; and SIDIAP-H in Spain), while three databases gave rates more than twice as high, at over 1,300/100,000 person-years (MDCD, MDCR, and OPTUM-SES in the US). Among women aged 75 to 84 years, the lowest incidence rate was 585/100,000 person-years (CPRD-GOLD in the UK) and the highest was 2,167/100,000 person-years (MDCR in the US). We could not find any consistent patterns to explain which databases yielded higher or lower rates across outcomes. Appendix Table 4 lists all database-specific incidence rates.

**Figure 1:**
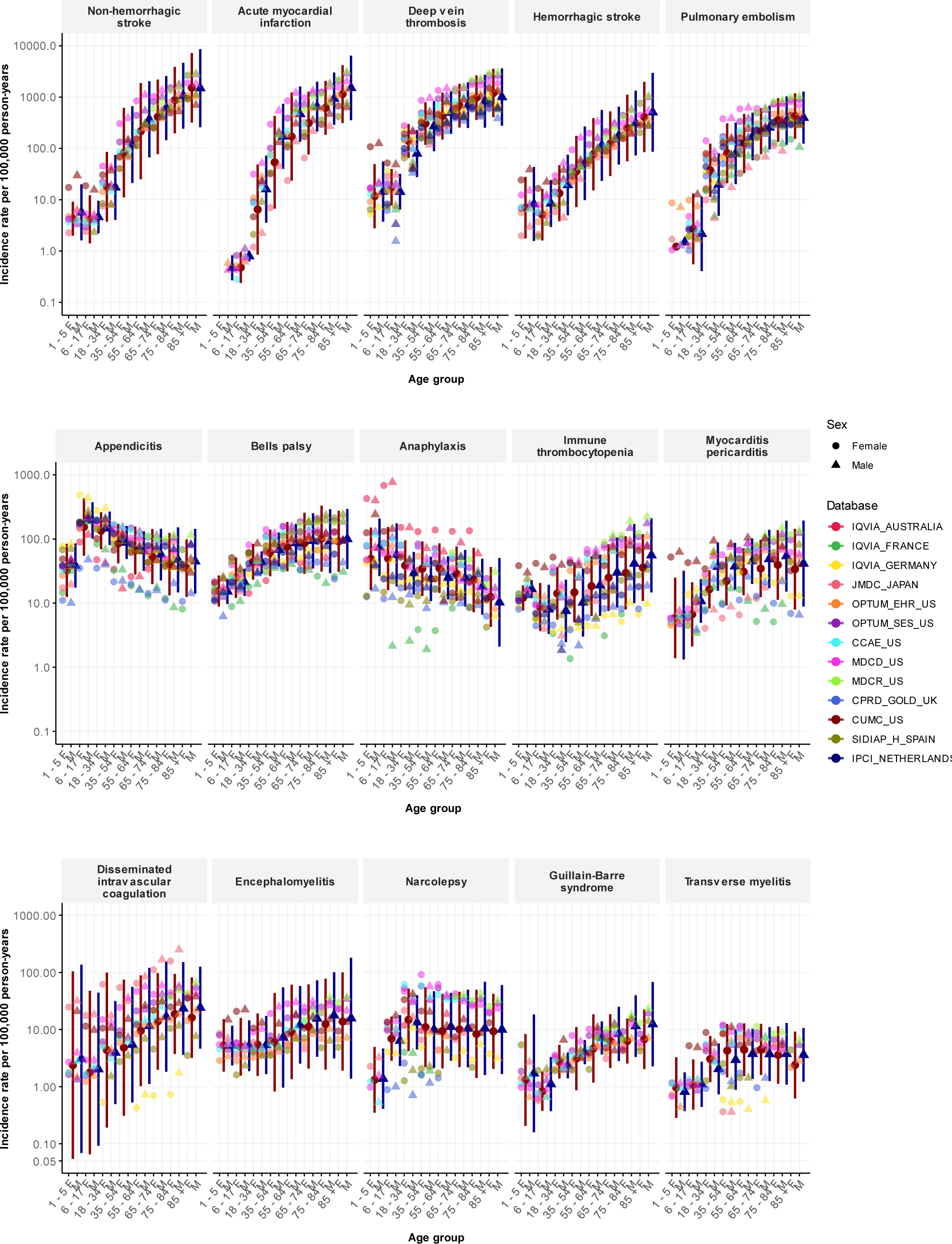
Age-sex stratified incidence rates, overall and per database, for 15 adverse events of special interest.

## Discussion

To our knowledge, this is the largest study to date on the descriptive epidemiology of the AESIs prioritised for post-marketing surveillance of COVID-19 vaccines. We report background rates of deep vein thrombosis, pulmonary embolism, stroke, immune thrombocytopenia, and disseminated intravascular coagulation, which are particularly relevance for COVID-19 vaccines given the observed effects of the SARS-COV-2 virus on coagulopathy.^14–17^

Our study provides a comprehensive, detailed assessment of the incidence rates of 15 AESIs across 13 databases, eight countries, and four continents. We found considerable heterogeneity between geographies and databases, suggesting that caution is needed when interpreting the difference between any observed and expected rates. We observed considerable variability with age and sex, emphasizing the need for standardization if background rates are used for surveillance purposes.

Many countries and organizations, such as the US and EU, have passive spontaneous adverse event reporting systems, such as the Vaccine Adverse Event Reporting System (VAERS). However, these systems cannot be used to calculate epidemiological estimates of the burden of disease, including incidence rates, because they lack denominators, which are the total number of persons or person-times being observed.^18,19^ Background incidence rates have been used to estimate the expected number of events in the general population.^3,19^ They are often obtained from the literature, but methods, case and population definitions, data, and calendar time can vary across publications. In contrast, we calculated all of the presented estimates using precisely the same setting and common analysis procedures, phenotyping algorithms, and data model. However, we still found substantial heterogeneity, suggesting that using a single overall estimate inadequately represents the true uncertainty around event incidence and may lead to confusion.

Population-level heterogeneity across data sources was substantial for all events, even after standardizing outcome definitions and stratifying by age and sex. While potentially concerning when seen together in one analysis, our findings are consistent with what has been observed in the literature. For example, in our study, the meta-analysis estimated incidence rates of transverse myelitis ranging from 1 to 4 per 100,000 person-years depend on age-sex strata. Previous studies have reported overall incidence rates of transverse myelitis ranging from 0·4 to 4·6 per 100,000 person-years.^19,20^ The rates of Bell’s palsy among those over 65 years old has ranged from 4·6 per 100,000 person-years in European data to 174 per 100,000 person-years in US data.^21,22^ Rates of narcolepsy from 1 to over 30 per 100,000 person-years have been reported among those aged 25 to 44 years.^21,22^ Recently reported data from the ACCESS project also showed heterogeneity in background rates.^5^ This heterogeneity needs to be taken into account when comparing rates across populations.

Most of the studied outcomes also had considerable within-source patient-level heterogeneity that followed age and sex patterns. For example, rates of cardiovascular diseases such as acute myocardial infarction, hemorrhagic and non-hemorrhagic stroke, deep vein thrombosis, and pulmonary embolism increased with age. The incidence of Guillain-Barre syndrome and Bell’s palsy also increased with age. Narcolepsy and appendicitis were more common in younger populations. The patterns observed in our study were generally comparable with previous reports.^2,3,21–25^ Stratification by age and sex or standardization are likely to be useful analytic strategies to reduce confounding when comparing incidence rates across populations. However, the magnitude of heterogeneity across sources within age-sex subgroups suggest that residual patient-level differences will remain, including the differences in the distributions of other risk factors such as comorbidities and medication use.

Comparing published results can be limited by differing study methodology, including the time-at-risk definition, study period, event definitions, population coverage, calendar year, and geographic location.^26^ Different subgroup definitions also make direct comparison difficult. For example, previous studies of Guillain-Barre syndrome have used age strata that do not fully overlap with each other.^2,5,21,25,27,28^ In contrast, because we applied the same definitions, data model, and analysis to all of the databases within our study, the heterogeneity observed cannot be attributed to analysis variability. Instead, it may have been due to differences in the underlying populations, healthcare systems, and data capture processes. Although some variability may have been due to systematic error, selection bias or differential outcome measurement error between databases, some may reflect true population differences such as socioeconomic status and comorbidities.

As we observed notable differences between age, sex, and databases, caution should be exercised when using incidence rates as a basis for comparison. Comparisons between incidence rates from different sources may be subject to substantial systematic error. We observed large variations between electronic health records and claims data sources when using the same analysis and outcome definitions. Comparisons with rates derived from randomised trials or spontaneous reporting data may have even greater variability. If observational databases are to be used to inform safety surveillance activities, within-database analyses (such as self-controlled case designs or propensity-score adjusted comparative cohort designs) may help reduce study bias for any given comparison. Demonstrating consistent effects across databases may further strengthen confidence in results. If observational data are used to derive historical ‘expected’ rates and compared against observed rates of events from another source, then the uncertainty in the background rate must be appropriately integrated to avoid misleading conclusions.

Our large number of participating databases, geographical coverage, and sizable study population allowed us to provide a comprehensive assessment of the background incidence rates across different healthcare systems and regions across the globe. Our study took advantage of the Observational Medical Outcomes Partnership common data model, which allowed us to use the same study design and analytical code and to gather results from participating data partners rapidly and without transferring patient-level data. All outcome definitions, clinical codes and phenotype algorithms have been made open source and are available online for review and to maximise reproducibility and reuse.

The primary limitation of this study is that all outcomes may have been subject to measurement error. As the outcome definitions were based on the presence of specific diagnostic codes and were not validated further, they may have had imperfect sensitivity or specificity. The analysis relied on data from 2017 to 2019 using a target population of all people in each database with >365 days of observation indexed on 1 January, 365 days’ time-at-risk, and outcome-specific clean windows to allow for recurrent events. The impact of these design decisions should be explored further.

## Conclusion

This study comprehensively assessed the descriptive epidemiology of potential AESIs for COVID-19 vaccines. It highlighted the wide range of adverse effects being monitored, from very rare neurological disorders to more common thromboembolic conditions. We reported large variations in the observable rates of AESI by age group and sex, demonstrating the need to account for stratification or standardization before using background rates for safety surveillance. We also found significant population-level heterogeneity in AESI rates between databases, implying that individual study estimates should be interpreted with caution and systematic error associated with database choice should be incorporated into any analysis. These background rates should provide useful real-world context to inform public health efforts aimed at ensuring patient safety while promoting the appropriate use of vaccines worldwide.

## Data Availability

Patient-level data cannot be shared without approval from data custodians due to local information governance and data protection regulations. Aggregated data, analytical code, and detailed definitions of algorithms for identifying the events are available in a GitHub repository (https://github.com/ohdsi-studies/Covid19VaccineAesiIncidenceCharacterization).

## Contributors

DAP, XL, PR, and GH conceived the idea of this study. XL, AO, GH, PR, and DAP were responsible for interpreting the results and writing the manuscript. RM, AZ, GR, and AS were responsible for implementing the study. XL, AO, PR, RM, KV, PRR, TDS, and MAS Contributed to the study execution (Data holders). XL, AO, GH, and PR Contributed to the study design. All the co-authors contributed to writing the manuscript. All authors approved the final version and had final responsibility for the decision to submit for publication.

## Declaration of interests

DPA’s research group has received research grants from the European Medicines Agency, from the Innovative Medicines Initiative, from Amgen, Chiesi, and from UCB Biopharma; and consultancy or speaker fees from Astellas, Amgen and UCB Biopharma. GH and AO receive funding from the US National Institutes of Health and the US Food and Drug Administration. KV and PR work for a research group who receives/received unconditional research grants from Yamanouchi, Pfizer-Boehringer Ingelheim, Novartis, GSK, Amgen, Chiesi none of which relates to the content of this paper. PBR, RM, AS, GR, AGS are employee of Janssen Research and Development, and shareholder in Johnson & Johnson. MAS receives grants and contracts from the US Food & Drug Administration and the US Department of Veterans Affairs within the scope of this research, and grants and contracts from the US National Institutes of Health, IQVIA and Private Health Management outside the scope of this research. Funders had no role in the conceptualization, design, data collection, analysis, decision to publish nor preparation of the manuscript. TDS has no conflicts of interest to declare. EM has no conflicts of interest to declare.

## Acknowledgement

This work was partially funded by the UK National Institute of Health Research (NIHR), European Medicines Agency, European Health Data & Evidence Network (EHDEN), US Food and Drug Administration CBER BEST Initiative (75F40120D00039), and US National Library of Medicine (R01 LM006910). EHDEN has received funding from the Innovative Medicines Initiative 2 Joint Undertaking (JU) under grant agreement No 806968. The JU receives support from the European Union’s Horizon 2020 research and innovation program and EFPIA.

DPA receives funding from NIHR in the form of a Senior Research Fellowship and the Oxford NIHR Biomedical Research Centre. XL receives funding from the Clarendon Fund (University of Oxford) to support her DPhil study.

We acknowledge English language editing by Dr Jennifer A. de Beyer, Centre for Statistics in Medicine, University of Oxford.

## Supplement materials

Appendix Table 1: database information

Appendix Table 2: adverse events of special interest phenotype definition and links

Appendix Table 3: source clinical codes used in phenotype definition

Appendix Table 4: age-sex specific crude incidence rates for each events of each database

## Notes

### Summary of Updates

Changes in references with new bibliography

